# Sex-specific neuroinflammatory responses to air pollution mediates cognitive performance in cognitively unimpaired individuals at risk of Alzheimer’s dementia

**DOI:** 10.1101/2024.08.09.24311733

**Authors:** Natalia Vilor-Tejedor, Blanca Rodríguez-Fernández, Patricia Genius, Alba Fernández-Bonet, Armand González-Escalante, Anna Brugulat-Serrat, Gonzalo Sanchez-Benavides, Irene Cumplido-Mayoral, Carolina Minguillón, Marta Cirach, Mark Nieuwenhuijsen, Karine Fauria, Gwendlyn Kollmorgen, Clara Quijano-Rubio, Henrik Zetterberg, Kaj Blennow, Arcadi Navarro, Marc Suárez-Calvet, Juan D. Gispert, the ALFA study

## Abstract

**Background:** A growing body of research links environmental factors with neurodegeneration and premature mortality. However, the biological mechanisms through which pollutants affect early Alzheimer’s disease (AD) pathology in asymptomatic stages are largely unknown. We aimed to assess the association between air pollution and changes in cerebrospinal fluid (CSF) biomarkers of AD and neuroinflammatory processes in cognitively unimpaired (CU) individuals at risk of AD dementia.

**Method:** We included 225 middle-aged CU participants from the ALFA+ study, many within the Alzheimer’s *continuum*, with baseline and one follow-up measurement of CSF biomarkers of primary pathology of AD (e.g. Aβ42/40 ratio, phosphorylated tau181 and total tau). Markers of neurodegeneration (e.g NfL), astrocytic (e.g. GFAP, YKL40), and microglial (e.g. sTREM 2) reactivity, and general inflammation (e.g. IL-6) were also included. Land use regression models were used to estimate individual levels of air pollution, including nitrogen oxide (NO_2_), particulate matter (PM_2.5_, PM_10_), and PM_2.5_ absorbance, at the participants’ residential address. We conducted sex-specific analyses using general linear models adjusted for age and time between measurements, with an interaction term for sex and environmental exposure. The influence of AD-related factors, like genetic predisposition and baseline amyloid pathology, on the relationship between air pollution and CSF biomarkers was also assessed. Moreover, we performed mediation analysis to investigate the pathways through which air pollution affected a 3-year rate of change in cognition and biological brain age via significant biomarkers.

**Results:** Results indicated sex-specific responses to air pollution. Women showed increases in IL-6 and GFAP, markers linked to neuroinflammation and astroglial activity, while men experienced impacts at baseline GFAP levels. The findings were consistent regardless of genetic predisposition to AD and amyloid pathology. Mediation analysis showed significant effects of GFAP on the relationship between air pollution and rate of change of attention and executive functions in women, highlighting primary influence pathways dependent on GFAP mediation. No significant mediation, neither direct effect was found for brain age.

**Conclusions:** Our findings highlight air pollution’s significant role in contributing to sex-specific neuroinflammatory and astrocytic response to air pollutants and its involvement in cognitive performance, underscoring the need for further research to elucidate these mechanisms.

**Highlights:**

● The study evaluates the 3-year rate of change in CSF biomarkers and incorporates an assessment of vulnerability based on genetic factors related to Alzheimer’s disease and amyloid pathology, enriching the understanding of individual differences in response to air pollution.
● The study establishes significant associations between air pollution and changes in CSF biomarkers related to neuroinflammatory processes in cognitively unimpaired individuals at risk of AD dementia.
● We identified distinct impacts of air pollution on men and women, with women showing more long term detrimental effects.
● CSF GFAP levels mediated the relationship between air pollution and rate of change of attention and executive functions in women.
● Mediation analysis showed that pathways through which air pollution affected a 3-year rate of change in cognition are significantly influenced by astrocytic reactivity.

## 1. Introduction

Air pollution, a complex mixture of solid particles and gases, is a recognized environmental hazard with substantial adverse health effects. Air pollution stands as the most significant environmental risk factor worldwide, and a major cause of premature death and disease, primarily due to its well-established links with respiratory and cardiovascular diseases (EEA, 2022). Recent studies have expanded our understanding of its impact, suggesting that air pollution extends beyond the respiratory and cardiovascular systems (Manisalidis et al., 2020), negatively affecting cognitive function and raising concerns about long-term implications for brain health (Chandra et al., 2022; Kioumourtzoglou et al., 2016; Power et al., 2016). Indeed, consistent evidence of an association between traffic-related air pollutants and clinical dementia was reported in a recent systematic review and meta-analysis (Wilker et al., 2023). For instance, a 2 to 3 μg/m^3^ increase in average yearly exposure to particulate matter with aerodynamic diameters equal to or less than 2.5 micrometers (PM_2.5_) was associated with a 4% increased risk of developing dementia. Our own earlier studies also revealed traffic-related air pollutants impact on cerebrospinal fluid (CSF) biomarkers and amyloid deposition (Alemany et al., 2021), cognition and brain gray and white matter volumes (Crous-Bou et al., 2020; Falcón et al., 2021). Furthermore, air pollution is now recognized as a modifiable risk factor for dementia, with potential prevention or delay of up to 3% of global dementia cases worldwide (Livingston et al., 2024). With an estimated 150 million people projected to have dementia by 2050, modifying air pollution exposure could prevent or delay the onset of dementia in approximately 3 million cases.

Numerous mechanisms have been proposed to elucidate the potential impact of air pollution on the brain and its role in the initiation of Alzheimer’s disease (AD). For instance, airborne pollutants may directly translocate to the cortex regions of the brain where the early stages of AD are believed to be initiated (Peters, 2023). Additionally, these pollutants can enter the bloodstream, eventually reaching the brain’s vasculature. In doing so, they can provoke neuronal inflammation and degeneration, potentially exacerbating the pathological processes associated with AD (Jankowska-Kieltyka et al., 2021). Furthermore, ultrafine particles found in the air can trigger inflammatory responses in various barrier organs. This systemic inflammation and the associated oxidative stress may collectively contribute to the progression of dementia and AD, underscoring the complex and multifaceted nature of the relationship between air pollution and neurodegenerative disorders (You et al., 2022).

Extensive research has shown the existence of sex-based disparities in the manifestation, diagnosis, and treatment responses related to AD (Ferretti et al., 2018). While environmental factors contribute to these differences (Bell et al., 2015), the specific mechanisms remain unclear, hindering the application of research findings. Understanding these mechanisms is crucial for enhancing AD risk assessment, as well as sex-specific prognosis, and treatment. Considering the previous evidence, this study aimed to evaluate sex-specific associations between traffic-related air pollutants and a three-year rate of change of AD core CSF, as well as other CSF biomarkers of neurodegeneration and neuroinflammation in cognitively unimpaired (CU) individuals at risk of AD. We explored whether the effects were independent of or mediated by genetic predisposition to AD and hallmarks of amyloid pathology. Finally, we evaluated whether significant biomarker alterations mediate the association between air pollution and a three-year rate of change of cognitive performance and biological brain age.

## 2. Material and methods

### 2.1 Study sample

The study sample comprised CU middle-aged individuals drawn from the ALFA+ cohort, a nested longitudinal study of the ALFA (for ALzheimer and Families) parent cohort (Molinuevo et al., 2016). The ALFA parent cohort was established as a research platform to understand the early pathophysiological alterations in preclinical AD and is composed of 2743 CU individuals (between 45 and 75 years) and enriched for family history of AD and genetic risk factors for AD (Vilor-Tejedor et al., 2024).

In the present study, we included 225 participants of the ALFA+ cohort, with available environmental exposure assessment, genotyping as well as CSF biomarkers collected at two distinct time point visits (the first from the baseline visit of the ALFA+ study [2016–2019], and the second from the visit 1 of the ALFA+ study [2019-2023]).

### 2.2 CSF Biomarkers Assessment

CSF biomarkers amyloidLβ (Aβ) 42, Aβ40, neurofilament light (NfL), soluble triggering receptor expressed on myeloid cells 2 (sTREM2), chitinase-3-like protein 1 (YKL40), glial fibrillary acidic protein (GFAP), and interleukin 6 (IL-6), were measured using the NeuroToolKit panel of immunoassays on either the cobas^®^ e 411 analyzer or cobas e 601 analyzer (Roche Diagnostic International Ltd, Rotkreuz, Switzerland. CSF phosphorylated tau181 (ptau181) and total tau (t-tau) were quantified using the electrochemiluminescence Elecsys^®^ Phospho-Tau (181P) CSF and Total-Tau CSF immunoassays, respectively, on the fully automated cobas e 601 analyzer (all Roche Diagnostics International Ltd., Rotkreuz, Switzerland), as previously described (Milà-Alomà et al., 2020). All fluid biomarkers were measured at the Clinical Neurochemistry Laboratory, Sahlgrenska University Hospital, Mölndal, Sweden.

### 2.3 Exposure Assessment

Air pollution measurements were collected as a part of the air pollution monitoring campaign of the European Study of Cohorts for Air Pollution Effects (ESCAPE). Following well-established protocols (Beelen et al., 2013; Eeftens et al., 2012), nitrogen oxide (NO_2_) and particulate matter (PM_2.5_ [particulate matter with aerodynamic diameter less than 2.5 μm], PM_10_ [less than 10 μm], and PM_2.5_ absorbance [PM_2.5_ light absorption]) were measured in three different seasons during 2009 (warm, cold, and one intermediate temperature season). Based on geographical information systems (GIS) and statistical methods, Land Use Regression models (LUR) were developed for the mentioned pollutants, representing the annual average for the reference year (2009). Individual levels of air pollution were estimated at the participant’s residential addresses reported in 2013-2014 representing a long-term exposure to air pollution. Further details can be found elsewhere (Alemany et al., 2021).

### 2.4 Cognitive measurements

Cognition was evaluated through a comprehensive neuropsychological battery covering the main cognitive domains: attention (WAIS-IV: Digit Span, WMS-IV: Symbol Span and TMT-A), episodic memory (Free and Cued Selective Reminding Test, Memory Binding Test, WMS-IV Logical Memory and NIH-toolbox Picture Sequence Memory test), and executive function (TMT-B, WAIS-IV Coding, WAIS-IV Matrix reasoning and NIH-toolbox Flanker Inhibition test). Cognitive change for each composite was computed by subtracting the follow-up visit score from the baseline cognitive score, representing negative values a worse performance in the follow-up visit.

### 2.5 Brain age estimation

Brain-age estimation was conducted using previously established brain-age prediction models (Cumplido-Mayoral et al., 2023). In brief, we employed XGBoost regressor models, separately trained for both women and men, utilizing the XGBoost Python package (https://xgboost.readthedocs.io/en/latest/python). These models were trained with input data comprising 183 FreeSurfer-derived regions for volumes and cortical thickness from the UK BioBank cohort. Subsequently, we applied this model to predict brain-age for the ALFA cohort, followed by the application of a well-established age-bias correction procedure (de Lange & Cole, 2020; Le et al., 2018). The bias-corrected predicted brain-age was then obtained by subtracting the individual’s chronological age, resulting in the calculation of the brain-age delta.

### 2.6 Genetic Profiling

DNA was obtained from blood samples through a salting out protocol. Genotyping was performed with the Illumina Infinium Neuro Consortium (NeuroChip) Array (build GRCh37/hg19) (Blauwendraat et al., 2017). Quality control procedure was performed using PLINK software. Imputation was performed using the Michigan Imputation Server with the haplotype Reference Consortium Panel (HRC r1.1 2016) (Das et al., 2016) following default parameters and established guidelines. Genetic predisposition to AD (PRS-AD) was calculated using the PRSice version 2 tool (Choi & O’Reilly, 2019). A full description of the genetic profiling of the ALFA study is available elsewhere (Vilor-Tejedor, Genius et al., 2024).

### 2.5 Statistical Analyses

Multiple linear regression models were used to evaluate the association between exposure to traffic-related air pollutants and both baseline levels and the rate of change in CSF biomarker levels. Sex-dependent effects were evaluated by including an interaction term in the models. Models were adjusted for age, sex (main models), *APOE*-ε4 status, and the time difference between acquisitions (follow-up models). To assess the robustness of our findings, several sensitivity analyses were performed, which included adjusting for potential confounding variables such as genetic predisposition to AD, CSF Aβ42/40, CSF ptau181/Aβ42, and CSF Aβ40 levels. Subgroup analyses were conducted to investigate whether the observed associations differed based on an individual’s genetic predisposition to AD. Finally, mediation models were assessed to investigate the pathways through which air pollution affected a 3-year rate of change in cognition and biological brain age via significant biomarkers. The mediation models analyzed the effects of air pollution on a 3 year rate of change on episodic memory, executive function and attention composites through changes in significant CSF biomarkers associated with levels of air pollution. Multiple comparisons corrections using the false discovery rate method were applied to control for potential type I errors in predefined clustering of biomarker families through a cluster analysis using the k-means method. The analyses were conducted using R version 4.3.2. The figures were generated with R version 4.3.2 and edited with Biorender.

## 3. Results

### 3.1 Descriptives

The study analyzed data from 225 participants, comprising 87 males and 138 females with complete information for all environmental predictors and CSF biomarkers. The average age was approximately 61 years, with no significant age difference observed between sexes. A higher percentage of men were *APOE*-ε4 carriers compared to women, although both groups exhibit a similar genetic risk profile for AD. No sex differences were found in CSF biomarker changes. Males generally were exposed to higher concentrations of NO_2_, PM_2.5_, and PM_2.5_ absorbance [**Table 1**]. Cross-correlation between CSF biomarkers and air pollutants is shown in **Supplementary Figure 1**]. A total of 5 optimal biomarker family groups were identified through cluster analysis to accurately adjust for multiple comparisons [**Supplementary Figure 2**].

**Table 1.** Main characteristics of the study population.

|  | Study population (N= 225) | Women (N=138) | Men (N=87) | p-value |
| --- | --- | --- | --- | --- |
| *Demographics* |  |  |  |  |
| Age at biomarkers assessment (years) (mean, SD) | 60.80 (4.58) | 60.60 (4.61) | 61.12 (4.55) | 0.410 |
| APOE status (N, %) |  |  |  | 0.040 |
| ε4 carriers | 119 (52.9%) | 65 (47.1%) | 54 (62.1%) |  |
| ε4 non-carriers | 106 (47.1%) | 73 (52.9%) | 33 (37.9%) |  |
| *Baseline biomarkers-CSF (log transformed mean, SD)* |  |  |  |  |
| Aβ42/40 ratio | 1.16 (0.67) | 1.17 (0.67) | 1.15 (0.67) | 0.243 |
| Aβ40 (pg/mL) | 4.24 (3.71) | 4.26 (3.70) | 4.21 (3.70) | 0.006 |
| p-tau181 (pg/mL) | 1.21 (0.82) | 1.22 (0.83) | 1.18 (0.78) | 0.090 |
| t-tau (pg/mL) | 2.29 (1.83) | 2.31 (1.84) | 2.27 (1.81) | 0.076 |
| NfL (pg/mL) | 1.91 (1.49) | 1.88 (1.37) | 1.97 (1.57) | <0.001 |
| GFAP (pg/mL) | 3.88 (3.36) | 3.86 (3.33) | 3.90 (3.38) | 0.024 |
| IL-6 (pg/mL) | 0.61 (0.34) | 0.62 (0.41) | 0.61 (0.16) | 0.778 |
| sTREM2 (pg/mL) | 3.90 (3.34) | 3.90 (3.32) | 3.91 (3.37) | 0.904 |
| *Rate of change biomarkers-CSF (log transformed mean, SD)* |  |  |  |  |
| Aβ42/40 ratio | 0.46 (0.57) | 0.46 (0.61) | 0.46 (0.49) | 0.921 |
| Aβ40 (ng/mL) | 2.83 (3.39) | 2.76 (3.43) | 2.92 (3.33) | 0.442 |
| p-tau181 (pg/mL) | 0.13 (0.78) | 0.17 (0.88) | 0.06 (0.29) | 0.652 |
| t-tau (pg/mL) | 1.22 (1.77) | 1.26 (1.86) | 1.14 (1.36) | 0.488 |
| NfL (pg/mL) | 1.33 (1.35) | 1.30 (1.26) | 1.39 (1.44) | 0.157 |
| GFAP (ng/mL) | 3.48 (3.16) | 3.47 (3.17) | 3.50 (3.13) | 0.158 |
| YKL40 (ng/mL) | 4.41 (4.34) | 4.43 (4.39) | 4.38 (4.24) | 0.281 |
| IL-6 (pg/mL) | -0.14 (0.53) | -0.66 (0.40) | 0.18 (0.64) | 0.013 |
| sTREM2 (ng/mL) | 3.20 (3.08) | 3.18 (3.13) | 3.24 (2.98) | 0.137 |
| *Air pollutants (mean, SD)* |  |  |  |  |
| NO2 (µg/m3) | 43.41 (17.97) | 39.92 (16.83) | 48.94 (18.43) | <0.001 |
| PM2.5 (µg/m3) | 15.37 (2.68) | 15.00 (2.55) | 15.95 (2.79) | 0.011 |
| PM10 (µg/m3) | 34.28 (6.18) | 33.25 (6.15) | 35.91 (5.91) | 0.001 |
| PM2.5 absorbance (µg/m3) | 2.29 (0.71) | 2.15 (0.62) | 2.52 (0.79) | <0.001 |

### 3.2 Sex-specific association of air pollution on CSF biomarkers levels at baseline

Results revealed significant sex-specific effects in the impact of PM_2.5_ absorbance and NO_2_ on CSF GFAP biomarker [**Supplemental Table 1**]. Additionally, the interaction between sex and PM_2.5_ showed a nominal significant association with GFAP. In all these models, higher levels were consistently observed in men compared to women [**Figure 1]**. Stratified models reveal that results were only significant in men, for which increased exposure to environmental pollutants corresponded with higher GFAP [**Figure 2]**. These findings were independent of genetic predisposition to AD, Aβ42/40 and ptau181/Aβ42 ratios and Aβ40. No significant associations were found in women at baseline, nor the whole sample.

**Figure 1.**
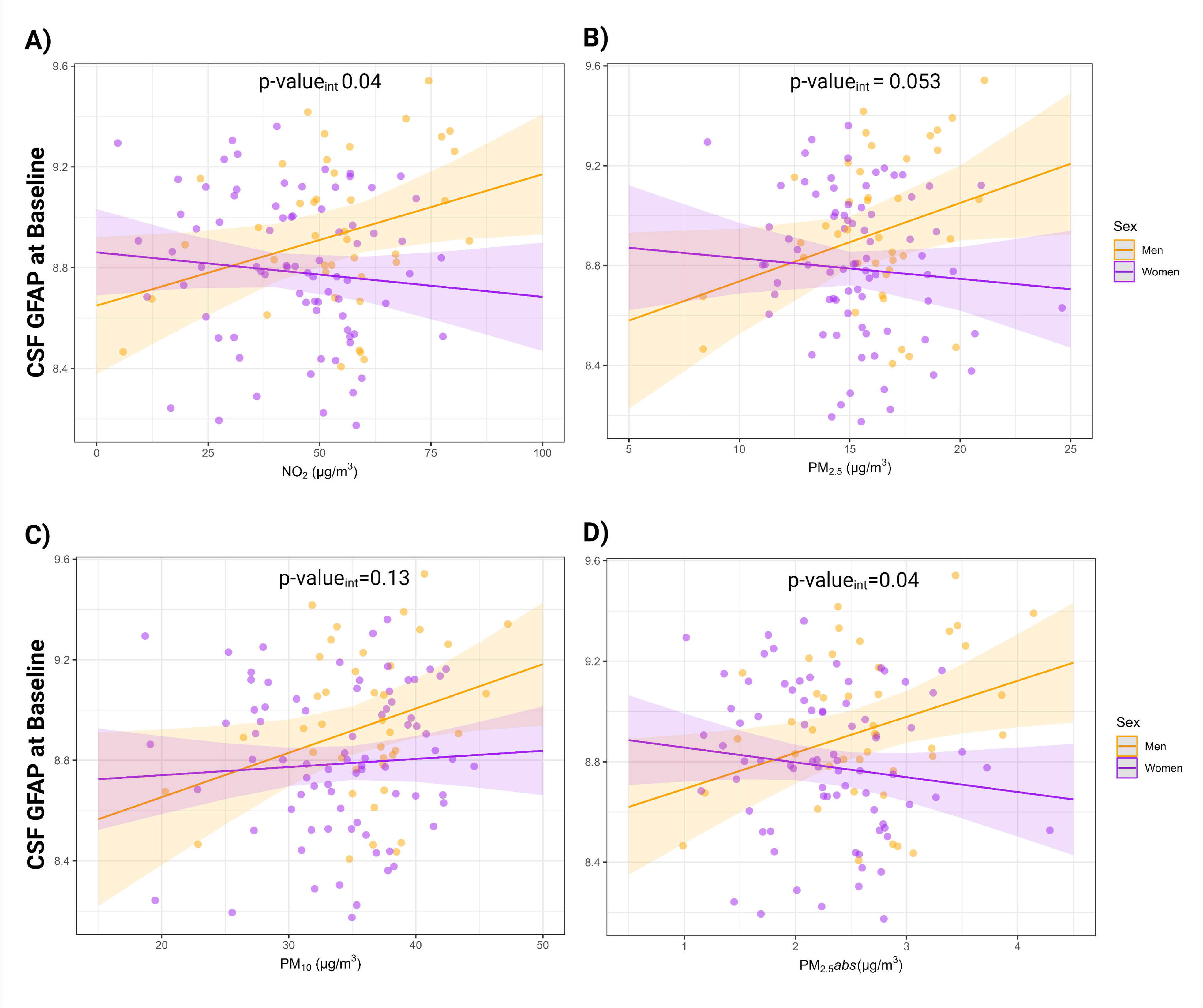
Significant interaction results between sex and air pollutants on CSF GFAP levels at baseline. A) Sex-specific interaction between Sex and NO_2_ B) Sex-specific interaction between Sex and PM_2.5_ C) Sex-specific interaction between Sex and PM_10_ D) Sex-specific interaction between Sex and PM_2.5abs._ *P-values for the interaction are presented after adjustment for multiple comparisons*.

**Figure 2.**
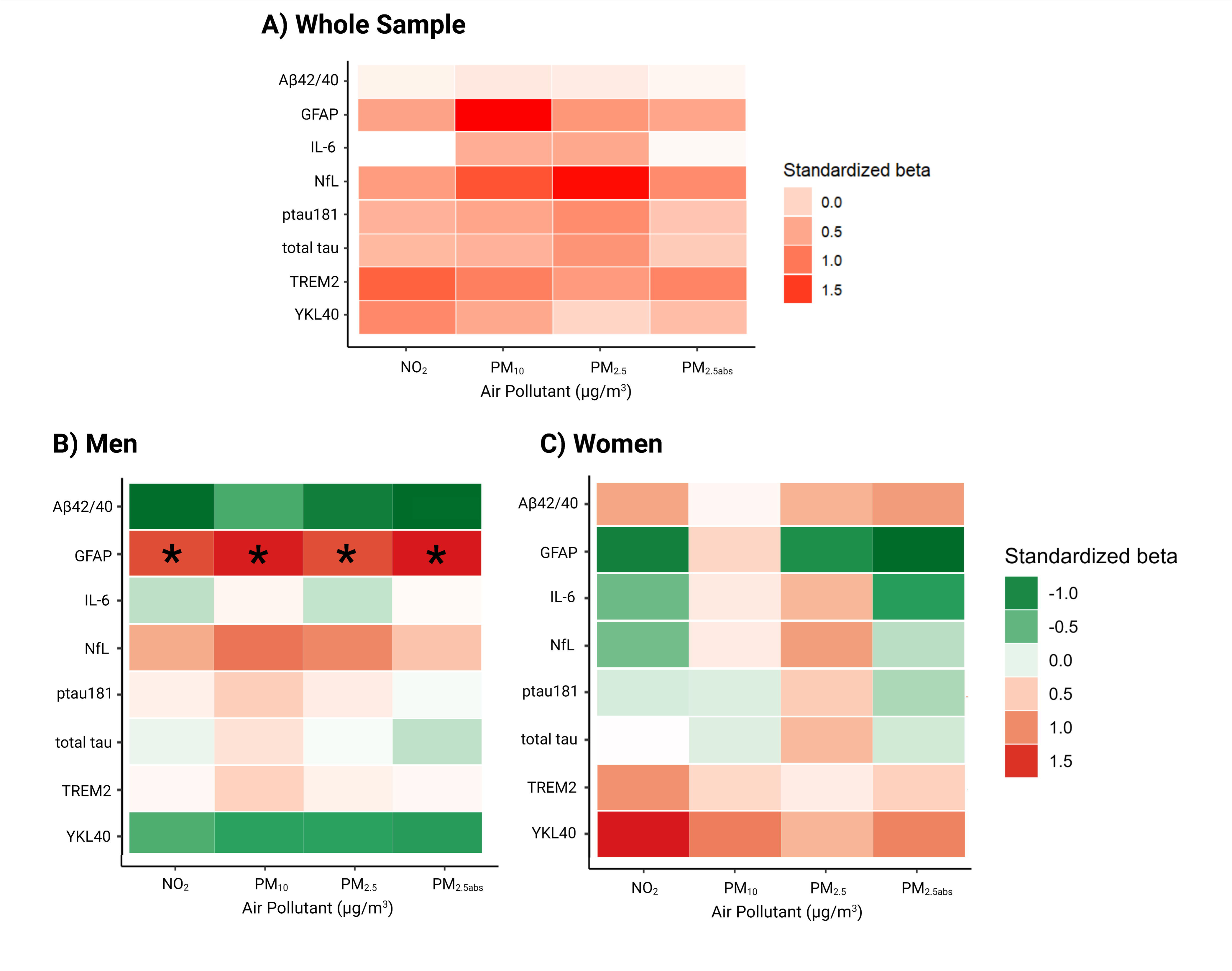
Main Effects and Sex-Stratified Models at Baseline. A) Results for the whole sample. B) Findings specific to men. C) Findings specific to women. Results that remain significant after adjusting for multiple testing are indicated. (*\*denotes findings where the False Discovery Rate is less than 0.05*).

### 3.3 Sex-specific association of air pollution and the rate of change of CSF biomarker

We observed a significant association between air pollution levels and 3-year rate of change in GFAP [**Figure 3, Supplemental Table 2-3**]. Specifically, higher exposure to NO_2_, was associated with 3-year increase in GFAP. Results highlighted sex-specific effects on the association between air pollution and changes in CSF. Notably, IL-6 exhibited a differential response to pollutants between sexes [**Figure 4, Supplemental Table 4**]. Stratified analyses by sex showed that these associations were exclusively significant in women. As exposure levels to NO_2_, PM_2.5_, PM_10_, and PM_2.5_ absorbance increased, a significant increase in IL-6 was observed in women, even at lower exposure levels [**Supplemental Table 5]**. All significant findings were independent of genetic predisposition to AD, CSF markers of amyloid pathology (Aβ42/40 ratio, and ptau181/Aβ40 ratio), and Aβ40. Diagnostic plots of significant models can be found in **Supplemental Figures 3 to 6**. To provide further insight into the robustness and assumptions of our models, we computed diagnostic plots of the significant models (see **Supplemental Figures 7 to 10**).

**Figure 3.**
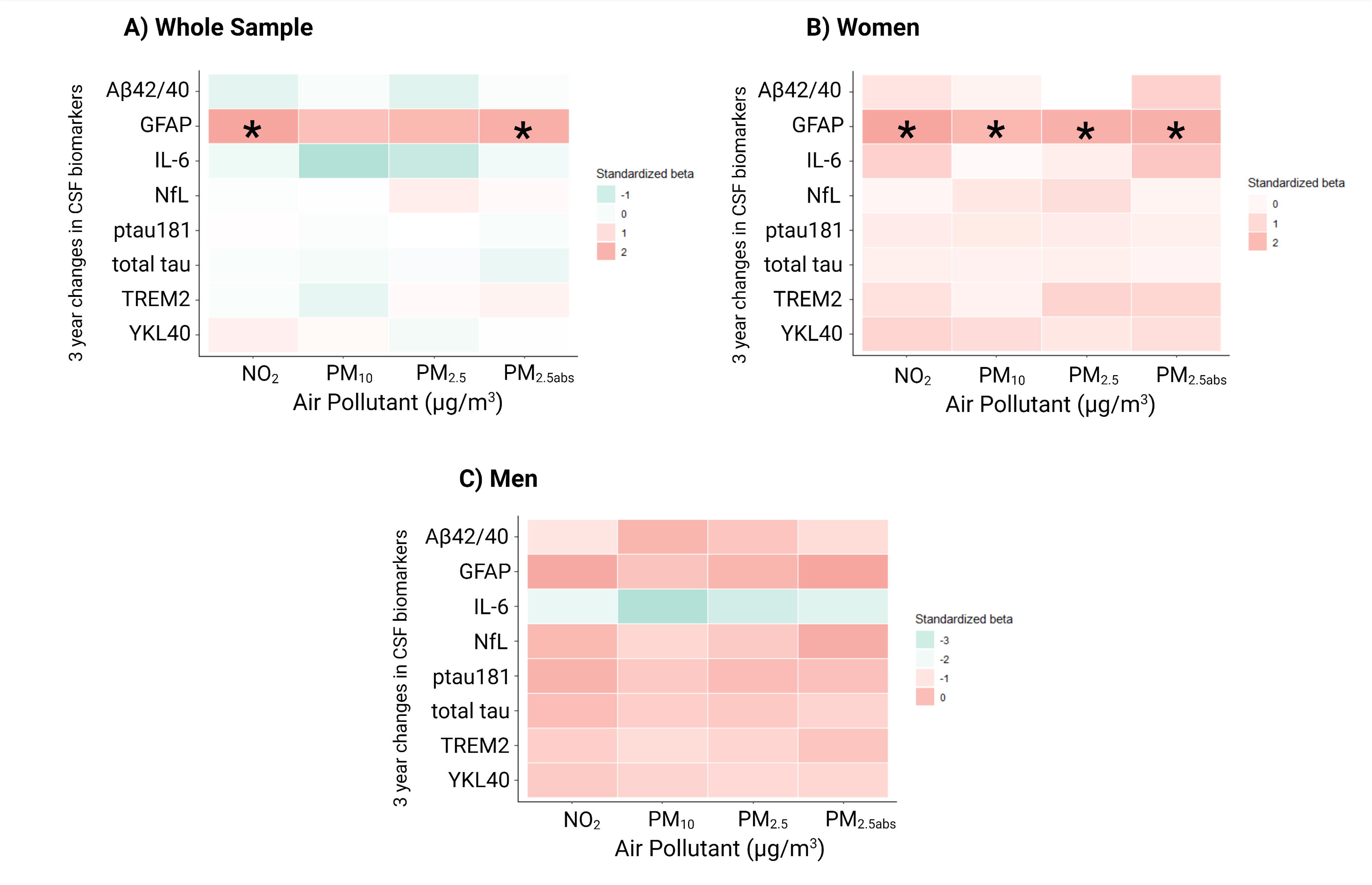
Association Between Air Pollution Levels and Rate of Change in CSF Biomarkers. A) Results for CSF biomarkers in the whole sample. B) Findings specific to CSF biomarkers in women. C) Findings specific to CSF biomarkers in men. (*\*denotes findings where the False Discovery Rate is less than 0.05*).

**Figure 4.**
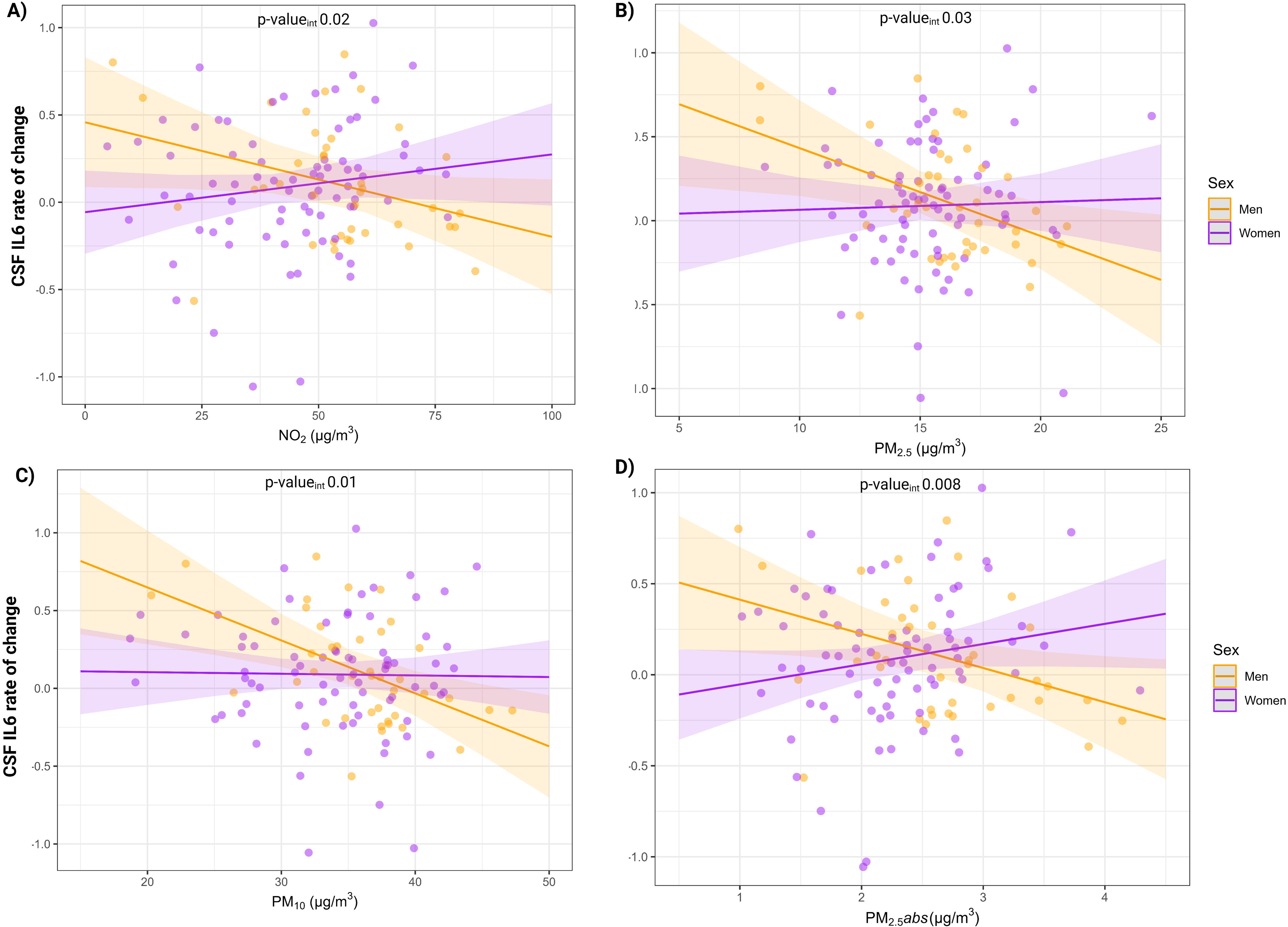
Sex-specific effects of air pollution on rate of change of biomarker levels. A) Significant interaction results between sex and air pollutants on CSF IL6 levels. *P-values for the interaction are presented after adjustment for multiple comparisons. X-axis levels are based on 2009 air pollution exposure models, used as a proxy for the data collection period. Reflects the stable spatial distribution of air pollution in Barcelona over the last 20 years*.

### 3.4 Mediation analysis of air pollution impact on the rate of change of cognitive domains

We explore the roles of significant biomarkers (IL-6, GFAP) in mediating the impact of air pollution exposure on 3-year rate of change in cognitive composites. The analysis revealed significant effects of GFAP on the relationship between PM_10_ levels and rate of change of attention and executive functions in women, highlighting primary influence pathways dependent on GFAP mediation. Moreover, a significant direct effect of PM_10_ was observed, indicating that increased levels of exposure to PM_10_ were associated with a reduction in attention scores, independent of biomarker alterations [**Figure 5**]. No significant direct, indirect, or total effects of air pollutants on biological brain age were found, even when considering the mediation roles of all biomarkers.

**Figure 5.**
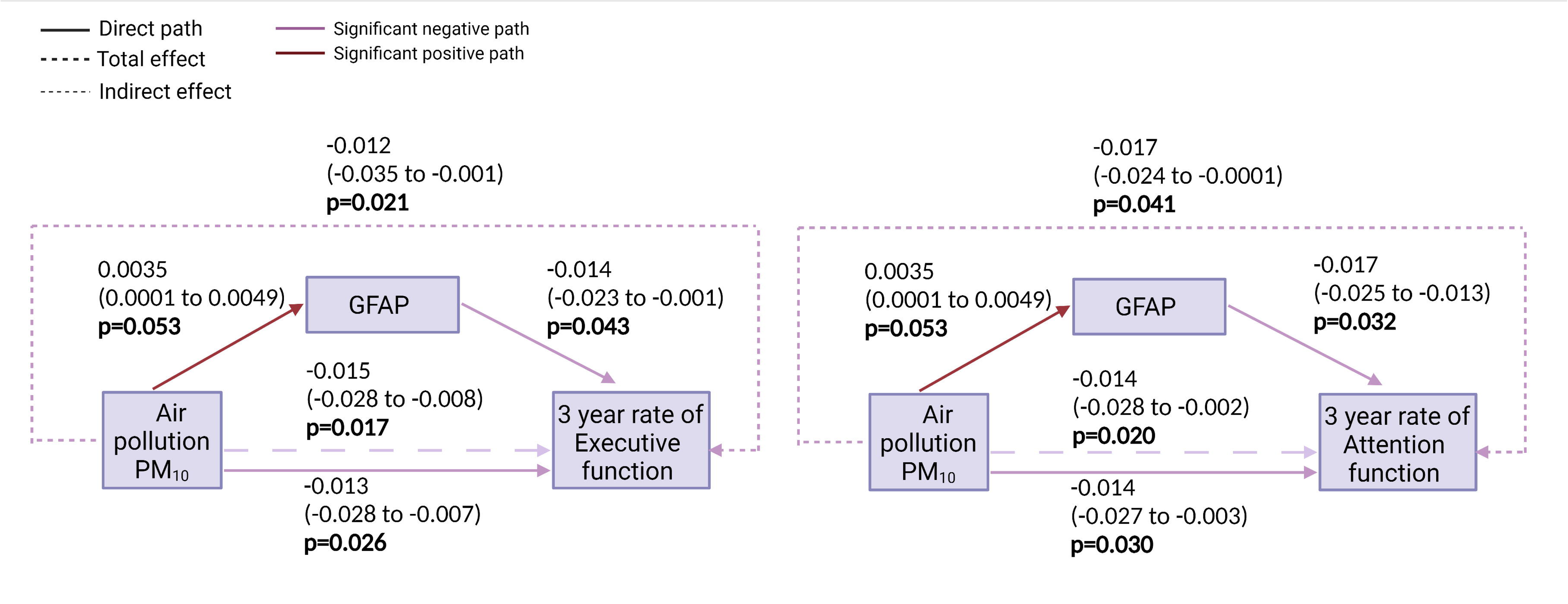
The mediating role of GFAP in the relationship between air pollution exposure and the rate of cognitive change in women.

## 4. Discussion

The current study examined the association between long-term air pollution levels, changes in CSF biomarkers and their impact in cognition, focusing on middle-aged CU individuals at risk of AD. Our findings showed significant relationships between air pollution exposure and three-year biomarker fluctuations, revealing notable differences based on sex. Specifically, results unveiled IL-6, as well as GFAP, as biomarkers that exhibit significant sex-specific changes in response to long-term air pollution exposure levels. The findings align with previous evidence implicating air pollution as a significant contributor to environmentally induced neuroinflammation in neurodegenerative processes (Block & Calderón-Garcidueñas, 2009; Brockmeyer & D’Angiulli, 2016; Jankowska-Kieltyka et al., 2021).

However, to the best of our knowledge, this is the first study to assess the relationship between environmental pollution and changes in CSF biomarkers related to amyloid pathology and neuroinflammation processes in cognitively unimpaired individuals at risk of developing AD. Moreover, our study extends this understanding by highlighting sex-specific variations in CSF biomarkers. Specifically, we observed elevated rates of change of GFAP in women as air pollution exposure levels increased. Increased CSF GFAP is typically indicative of astrocytic reactivity, a response commonly observed in the presence of various brain injuries and pathological conditions. Previous research has consistently demonstrated that air pollution drives microglia-astrocyte crosstalk, leading to their proinflammatory activation and the induction of oxidative stress (Song et al., 2022). Additionally, glial priming due to exposure to air pollution has been linked to neural dysfunctions, deficits in neurodevelopment, and the promotion of neurodegeneration (Gómez-Budia et al., 2020). Astrocytic reactivity begins from the pre-symptomatic stage of AD and is associated with brain Aβ load (Benedet et al., 2021; Chatterjee et al., 2021). Our findings reinforce the role of pollutants in triggering neuroinflammatory responses and subsequent astrocyte activation (Block & Calderón-Garcidueñas, 2009) with detrimental effects in women, which has been suggested to be associated with AD (Kim et al., 2023). Furthermore, as the level of exposure to air pollution increased, we observed differential levels of CSF IL-6 between sex, with a particular emphasis on the adverse effects observed in women. IL-6 plays a pivotal role as a key mediator of the immune response and neuroinflammation within the brain. Elevated CSF IL-6 signifies the presence of neuroinflammation, implying that exposure to pollutants may trigger or contribute to neuroinflammation in women. While no previous studies have reported the association between changes in CSF IL-6 and levels of air pollution in adults, especially when assessing sex-specific effects, a few studies have reported associations between traffic-related air pollution and neuroinflammation in children and young adults (Calderón-Garcidueñas et al., 2008, 2020).

Moreover, mediation analysis in our study showed significant effects of GFAP on the relationship between air pollution and rate of change of attention and executive functions in women, highlighting primary influence pathways dependent on astroglia reactivity mediation. We also observed a direct significant effect of air pollution on cognitive performance. These findings suggest that through this mechanism, air pollution might expedite the average age of dementia development. However, the absence of significant direct, indirect and mediated findings for biological brain age emphasizes the need of further research to fully understand the implications (Gunawan et al., 2024; Shi et al., 2021). This may indicate that the neurobiological mechanisms affecting brain age differ from those influencing more specific cognitive functions. Brain age may be less directly affected by air pollution variables or may involve different biological or environmental factors not accounted for in the current analysis.

Alternatively, other pathways might be responsible for mediating the effects of air pollution on brain age, suggesting the need for a broader exploration of potential mediators or different types of pollutants. Additional limitations should be considered when interpreting the findings of our study. First, our sample was specifically chosen to include individuals at risk for AD dementia, and as a result, generalizations to the broader population should be made with caution. Notably, these individuals have a higher genetic predisposition to AD compared to the general population and many of them show abnormal levels of AD biomarkers. However, it is important to highlight that our analyses were adjusted for genetic predisposition to AD in order to account for this variation and potential effect. Second, our estimations of air pollution exposure primarily relied on participants’ residential addresses, therefore, may not accurately reflect individual exposure levels. Unfortunately, we did not have access to data related to mobility or workplace exposure, which will be quantified and included in future studies to reduce the potential for exposure misclassification. Lastly, we used air pollution exposure data from 2009 models as a surrogate for the period of data collection, which occurred 4–10 years later. Nevertheless, it is worth noting that evidence suggests the spatial distribution of air pollution in Barcelona has remained relatively consistent over the past two decades (Cesaroni et al., 2013; Wang et al., 2013).

Nevertheless, our findings are reinforced by a consistent pattern of results, and underscore the emerging significance of air pollution as a prominent environmental risk factor for AD dementia. For instance, the observed associations between air pollutants and CSF neuroinflammatory biomarkers and their influence in cognition showed a potential contribution to the etiopathogenesis of AD dementia through neuroinflammatory mechanisms. Given the widespread prevalence of these exposures, even if the associated risk is of modest effect size, reductions in population-level exposure hold the potential to reduce the incidence of the disease. Moreover, future analyses within the longitudinal ALFA+ study and considering data related to mobility or workplace exposure will enable us to further explore whether air pollution effects are linked to disease progression, which is essential to elucidate the underlying mechanisms of the disease. In addition, the implications of these findings for research on neurodegeneration/dementia are significant. They suggest that environmental factors, such as air pollution, should be considered in the early assessment and potential intervention strategies for dementia prevention, targeting specific vulnerable groups of individuals. Future research should aim to replicate these findings in larger and more diverse cohorts. Additionally, longitudinal studies tracking these biomarkers over an extended period could provide further insight into the clinical progression to AD in relation to environmental factors. Understanding the molecular pathways underlying these observations would also be valuable, potentially revealing new targets for therapeutic intervention.

## 5. Conclusion

In summary, our study enhances our comprehension of the intricate relationship between air pollution and neuroinflammation in individuals at risk of AD, shedding light on the role of environmental factors in neuroinflammatory processes potentially preceding the development of cognitive decline and AD dementia. Furthermore, these findings imply that air pollution may be a relevant factor contributing to sex-specific differences in neurodegeneration associated with neuroinflammation, indicating in women a broader impact on astroglial reactivity, leading to their proinflammatory activation and the induction of oxidative stress. These findings also showed a mediatory impact on the 3-year rate of change of attention and executive functions exclusively in women, reinforcing the importance of considering biological sex and gender factors as relevant factors in studies of environmental factors affecting cognitive health and AD dementia.

These results advocate for equitable public health initiatives aimed at reducing the impact of air pollution on cognitive health.

## Data Availability

All data produced in the present study are available upon reasonable request to the authors.

## Conflicts of Interest

GK is a fullLtime employee of Roche Diagnostics GmbH, Penzberg, Germany. CQ-R is a full-time employee of Roche Diagnostics International Ltd, Rotkreuz, Switzerland. MS-C has given lectures in symposia sponsored by Almirall, Eli Lilly, Novo Nordisk, Roche Diagnostics, and Roche Farma; received consultancy fees (paid to the institution) from Roche Diagnostics; and served on advisory boards of Roche Diagnostics and Grifols. He was granted a project and is a site investigator of a clinical trial (funded to the institution) by Roche Diagnostics. In-kind support for research (to the institution) was received from ADx Neurosciences, Alamar Biosciences, Avid Radiopharmaceuticals, Eli Lilly, Fujirebio, Janssen Research & Development, and Roche Diagnostics. JDG has received research support from GE Healthcare, Roche Diagnostics, and Hoffmann – La Roche, speaker/consulting fees by Biogen, Roche Diagnostics, Philips Nederlands and Life Molecular Imaging, and serves in the Molecular Neuroimaging Scientific Advisory Board of Prothena Biosciences. HZ has served at scientific advisory boards and/or as a consultant for Abbvie, Acumen, Alector, Alzinova, ALZPath, Amylyx, Annexon, Apellis, Artery Therapeutics, AZTherapies, Cognito Therapeutics, CogRx, Denali, Eisai, Merry Life, Nervgen, Novo Nordisk, Optoceutics, Passage Bio, Pinteon Therapeutics, Prothena, Red Abbey Labs, reMYND, Roche, Samumed, Siemens Healthineers, Triplet Therapeutics, and Wave, has given lectures in symposia sponsored by Alzecure, Biogen, Cellectricon, Fujirebio, Lilly, Novo Nordisk, and Roche, and is a co-founder of Brain Biomarker Solutions in Gothenburg AB (BBS), which is a part of the GU Ventures Incubator Program (outside submitted work).

The NeuroToolKit is a panel of exploratory prototype assays designed to robustly evaluate biomarkers associated with key pathologic events characteristic of AD and other neurological disorders, used for research purposes only and not approved for clinical use (Roche Diagnostics International Ltd, Rotkreuz, Switzerland). Elecsys Phospho-Tau (181P) CSF and Elecsys Total-Tau CSF assays are approved for clinical use. COBAS and ELECSYS are trademarks of Roche. All other product names and trademarks are the property of their respective owners.’

## Acknowledgements

This publication is part of the ALFA study (ALzheimer’s and FAmilies). The authors would like to express their most sincere gratitude to the ALFA project participants, without whom this research would have not been possible. The authors would like to thank Roche Diagnostics International Ltd. for kindly providing the kits for the CSF analysis of study participants.

Collaborators of the ALFA study are: Federica Anastasi, Annabella Beteta, Lidia Canals, Alba Cañas, Marta del Campo, Carme Deulofeu, Maria Emilio, Carme Deulofeu, Ruth Dominguez, Maria Emilio, Sherezade Fuentes, Oriol Grau-Rivera, Laura Hernández, Felipe Hernández, Gema Huesa, Jordi Huguet, Laura Iglesias, Esther Jiménez, David López-Martos, Paula Marne, Tania Menchón, Paula Ortiz-Romero, Wiesje Pelkmans, Albina Polo, Sandra Pradas, Mahnaz Shekari, Lluís Solsona, Anna Soteras, Núria Tort-Colet, and Marc Vilanova.

## Supplementary Materials

**Supplemental Table 1**. Association of air pollution with CSF biomarkers at Baseline.

**Supplemental Table 2**. Associations of air pollution with 3 years rate of change in CSF biomarkers.

**Supplemental Table 3.** Sensitivity analyses of significant models analyzing the association between air pollution and rate of change in CSF biomarker levels.

**Supplemental Table 4.** Sex-Interaction effects on the association between air pollution and rate of change in CSF biomarker levels.

**Supplemental Table 5.** Significant models analyzing the association between air pollution and the rate of change in CSF biomarker levels in women.

**Supplemental Figure 1**. Cross-Correlation. A) CSF biomarkers at baseline. B) CSF biomarkers, 3 years rate of change. C) Air Pollutants.

**Supplemental Figure 2**. Optimal number of biomarker clusters for multiple comparisons correction.

**Supplemental Figures 3-6.** Performance of the CSF-GFAP model in assessing the significance of air pollution exposure across individuals.

**Supplemental Figures 7-10.** Performance of the CSF-IL-6 model in assessing the significance of air pollution exposure across individuals.

**E-mail address co-authors:**

Natalia Vilor-Tejedor

Blanca Rodríguez-Fernández

Patricia Genius

Maria Alba Fernández

Armand González-Escalante

Anna Brugulat-Serrat

Gonzalo Sanchez-Benavides

Irene Cumplido-Mayoral

Carolina Minguillón

Marta Cirach

Mark Nieuwenhuijsen

Karine Fauria

Gwendlyn Kollmorgen

Clara Quijano-Rubio

Henrik Zetterberg

Kaj Blennow

Arcadi Navarro

Marc Suárez-Calvet

Juan D. Gispert

