## Supplementary figures and images for "Sex-specific neuroinflammatory responses to air pollution mediates cognitive performance in cognitively unimpaired individuals at risk of Alzheimer’s dementia"

### SupplFig1.png

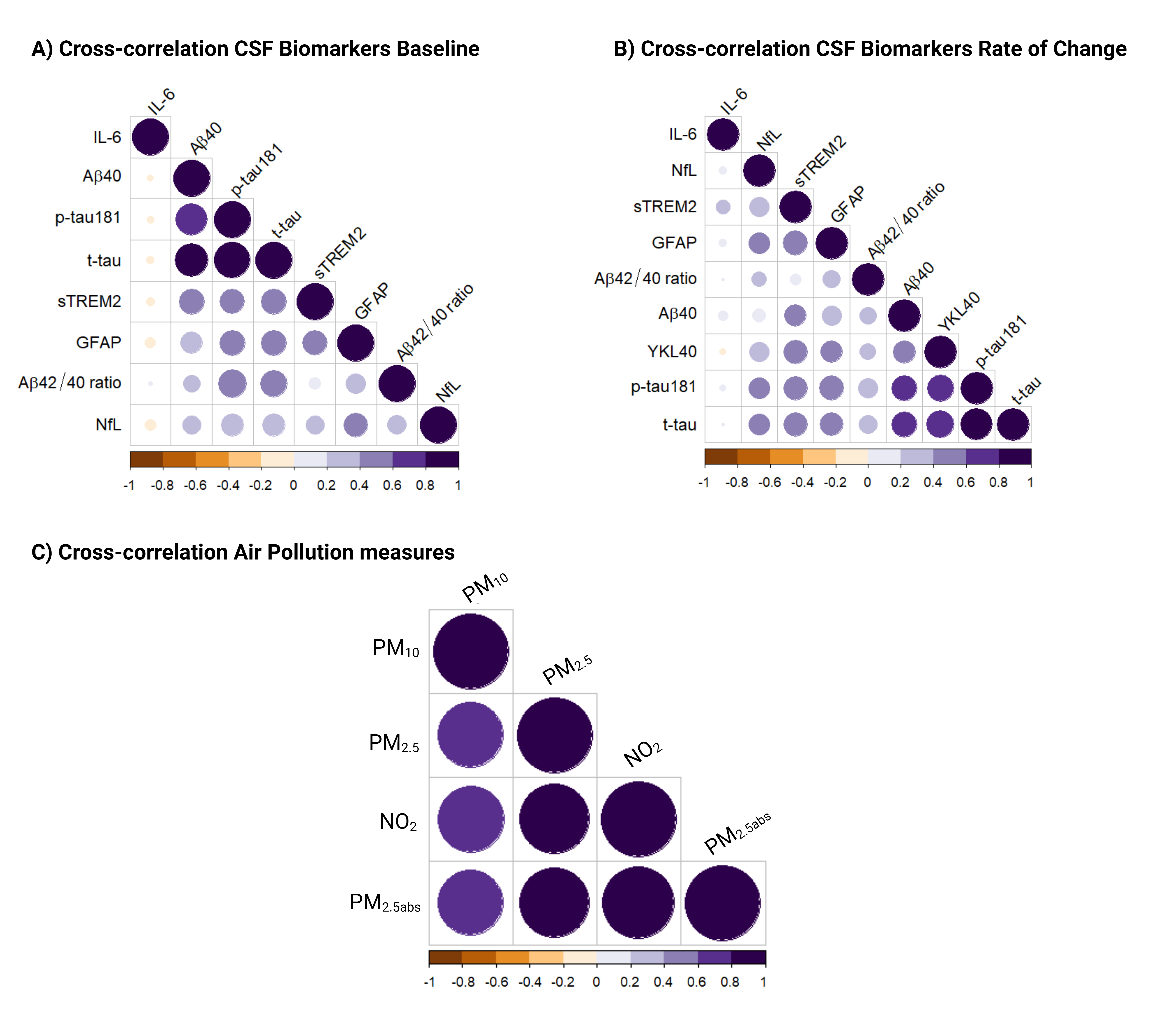

### SupplFig2NEW.png

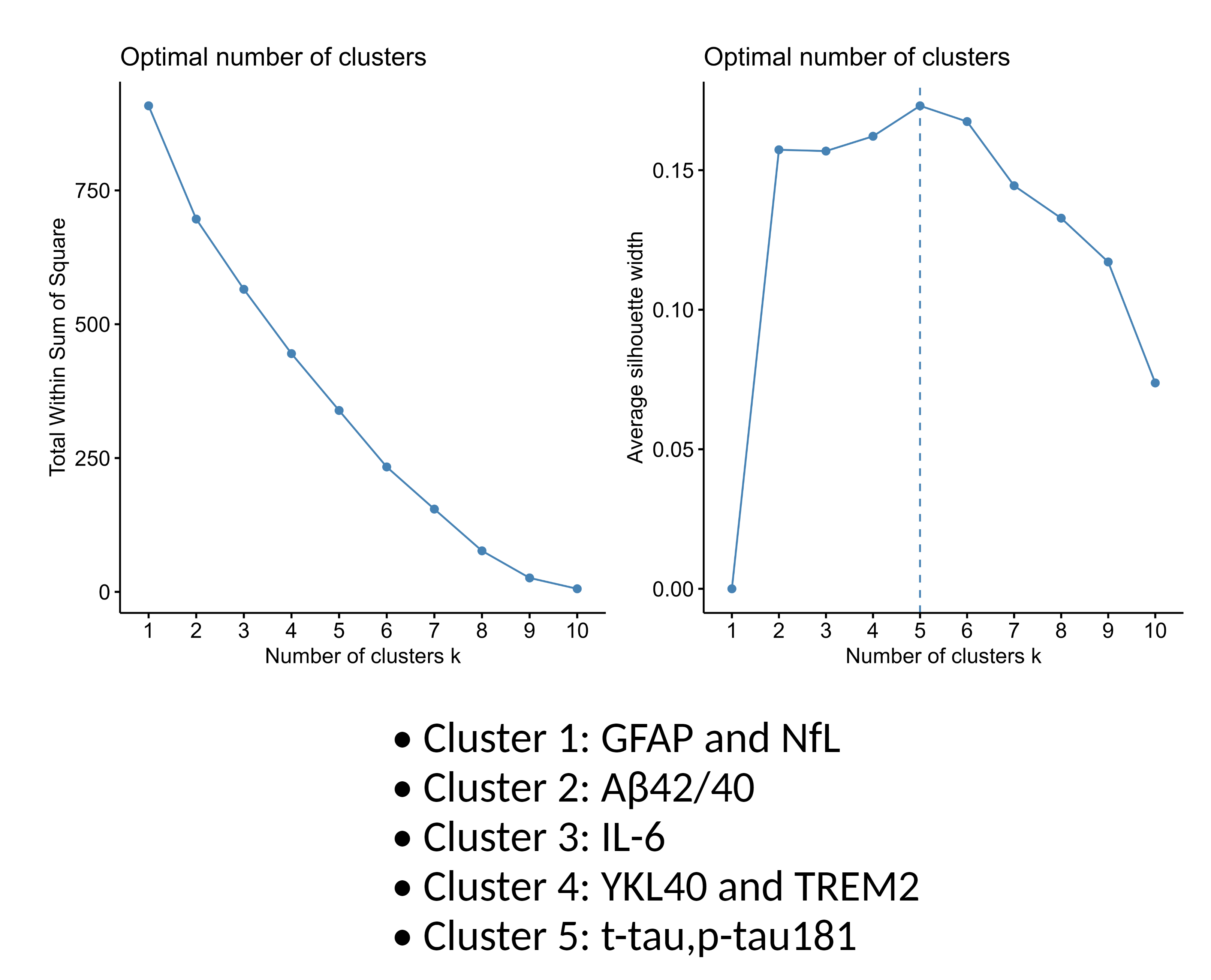

### SupplFig3-ModelPerformance_CSFGFAP_NO2_AllIndividuals_LogTransform.png

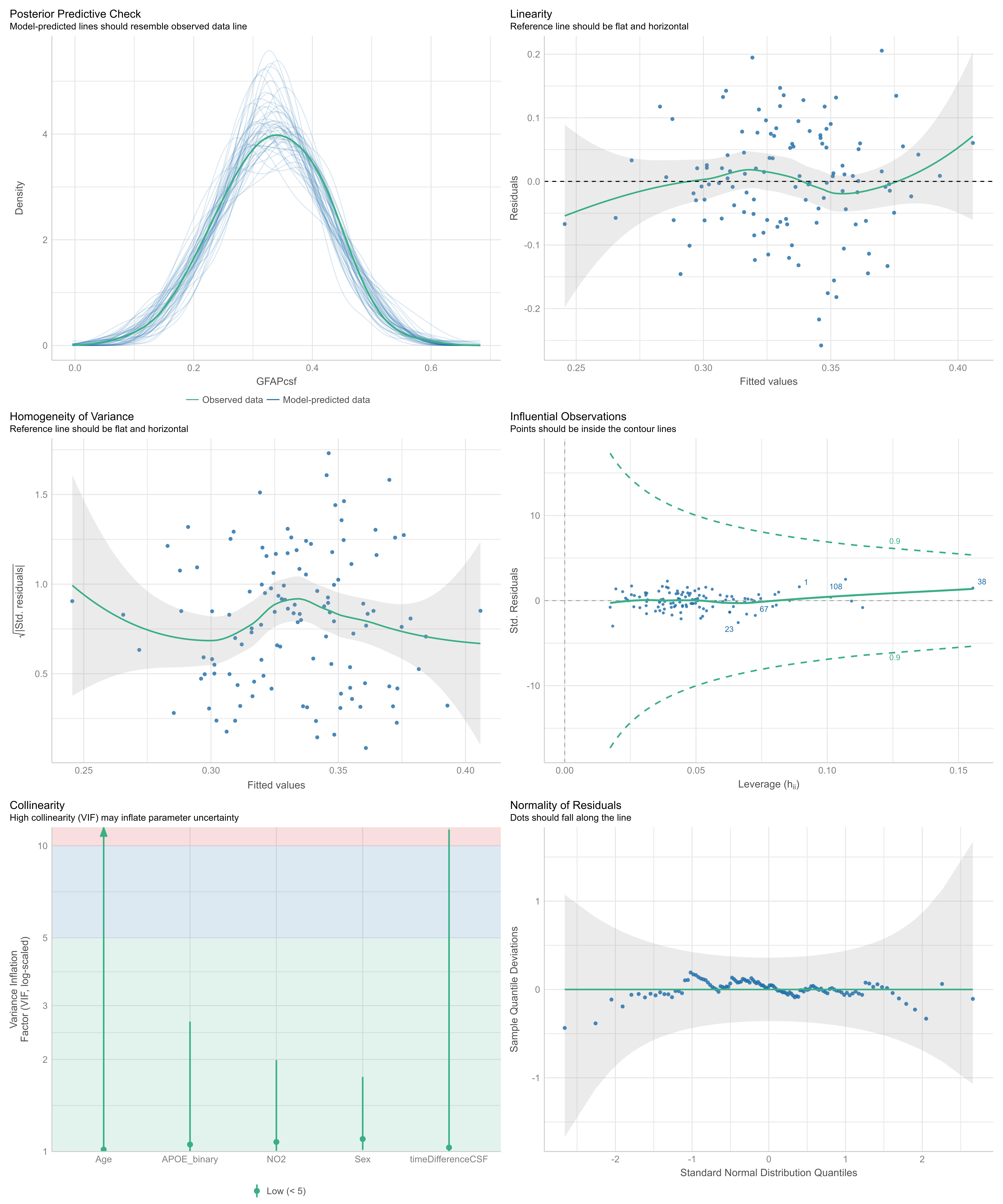

### SupplFig4-ModelPerformance_CSFGFAP_PM10_AllIndividuals_LogTransform.png

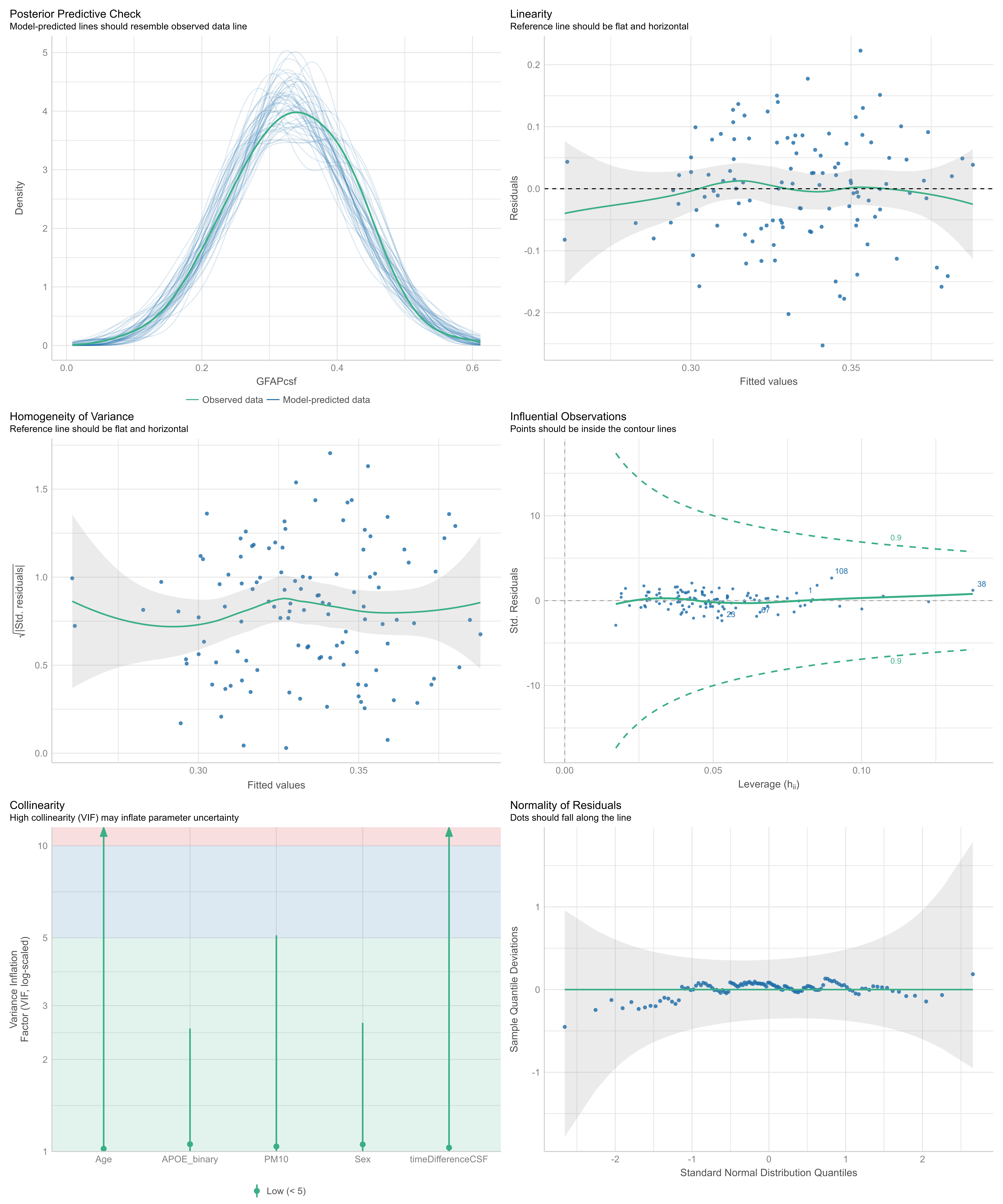

### SupplFig5-ModelPerformance_CSFGFAP_PM25_AllIndividuals_LogTransform.png

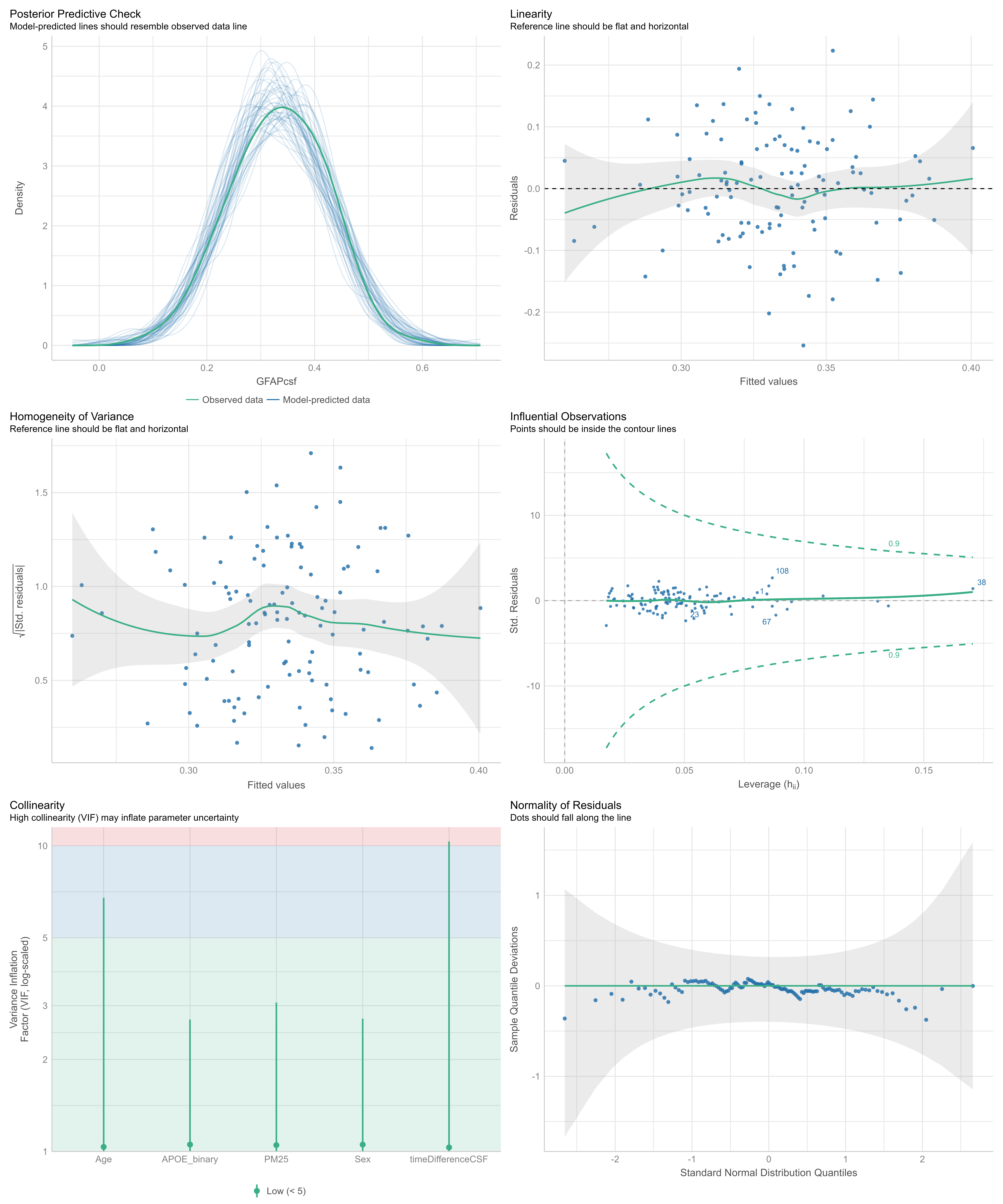

### SupplFig6-ModelPerformance_CSFGFAP_PM25abs_AllIndividuals_LogTransform.png

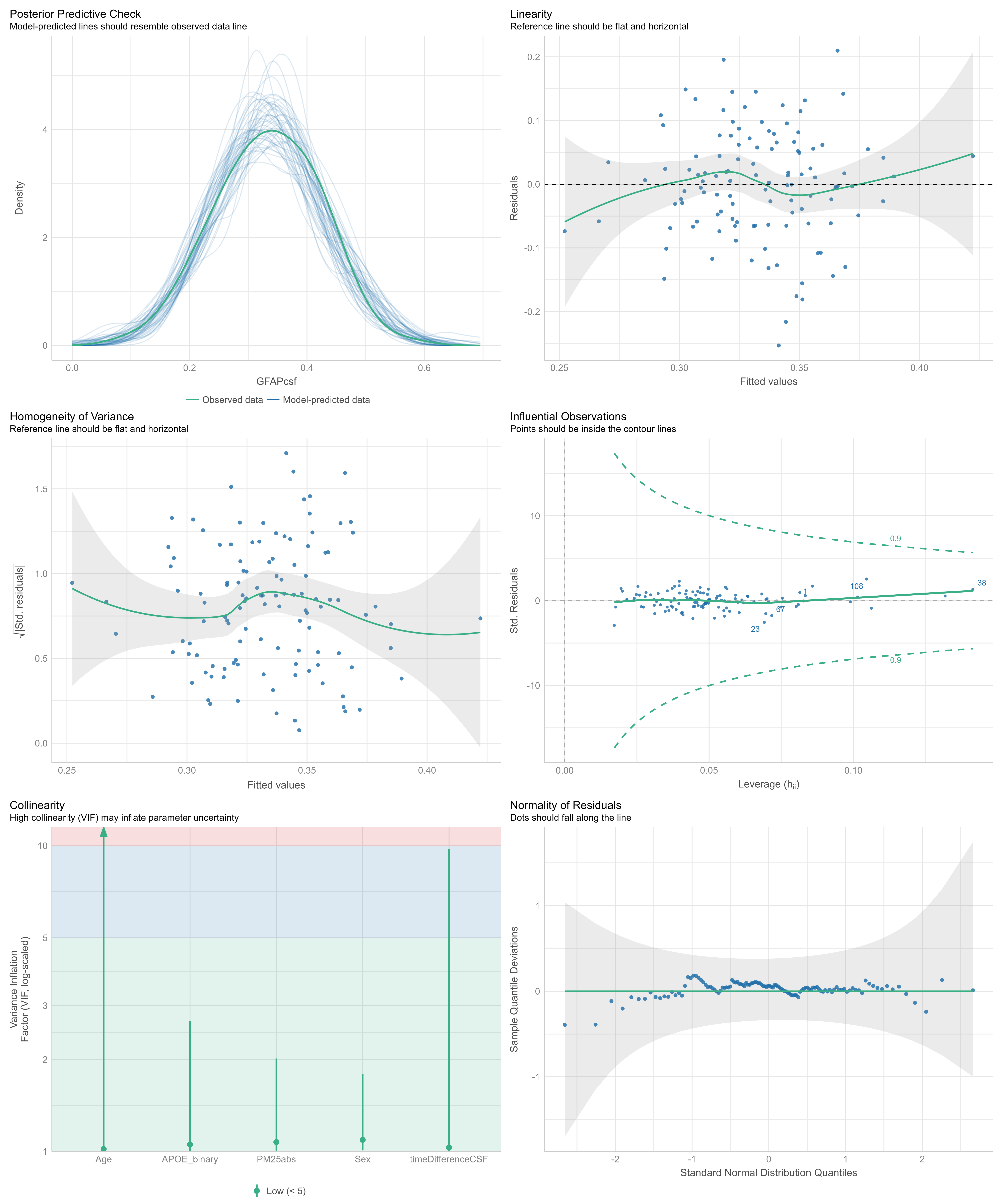

### SupplFigure7-ModelPerformance_CSFIL6_NO2.png

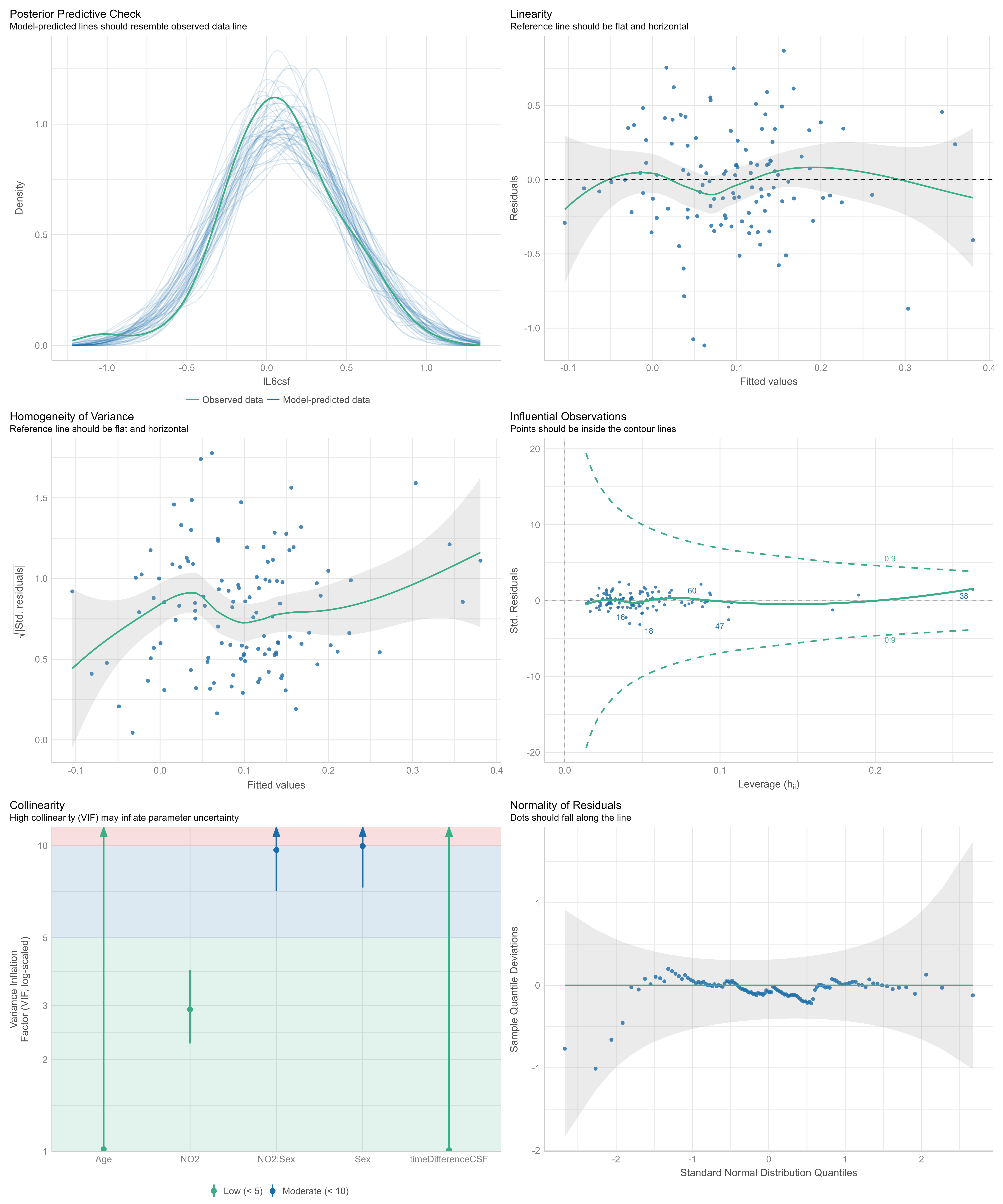

### SupplFigure8-ModelPerformance_CSFIL6__PM10.png

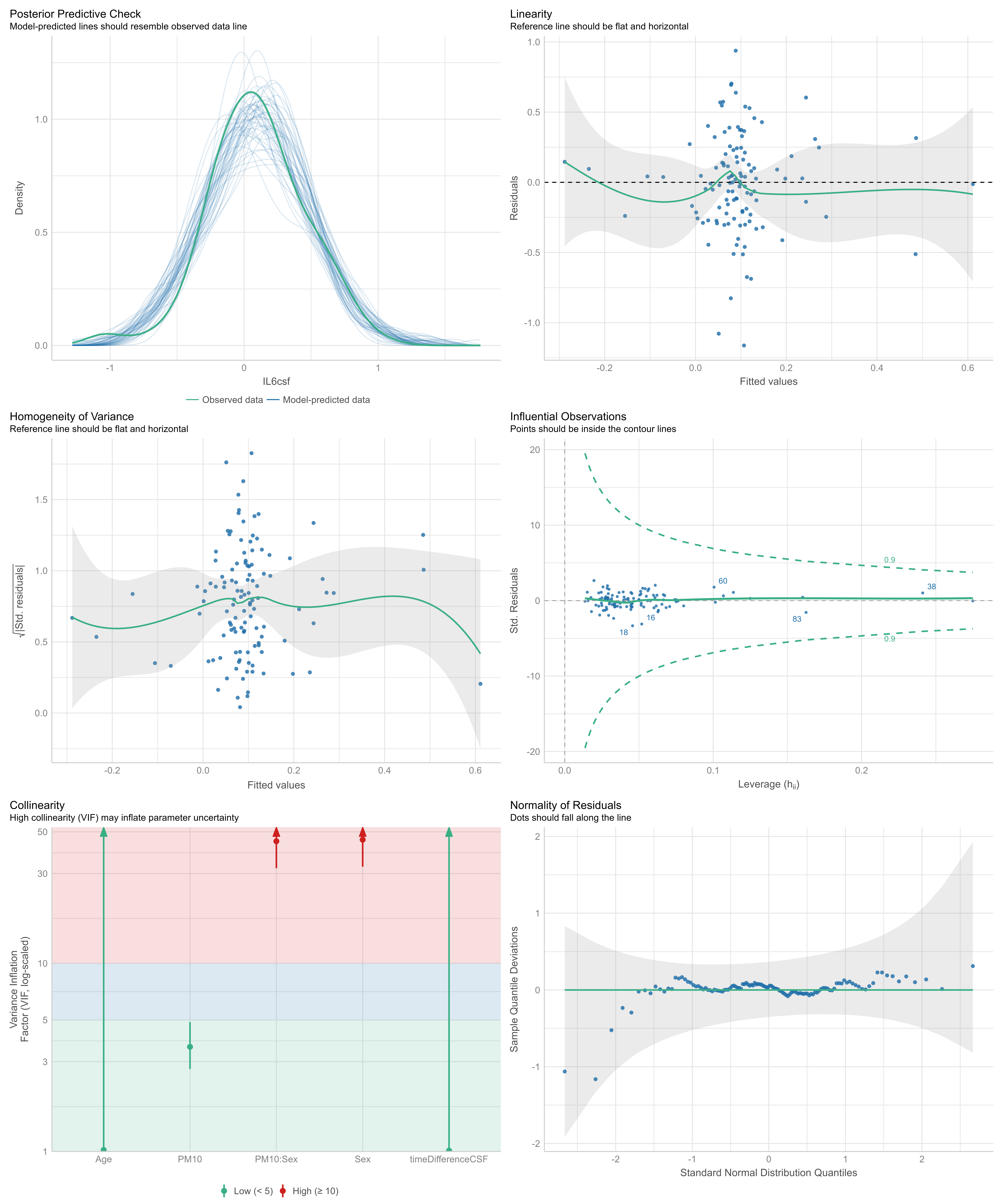

### SupplFigure9-ModelPerformance_CSFIL6_PM25.png

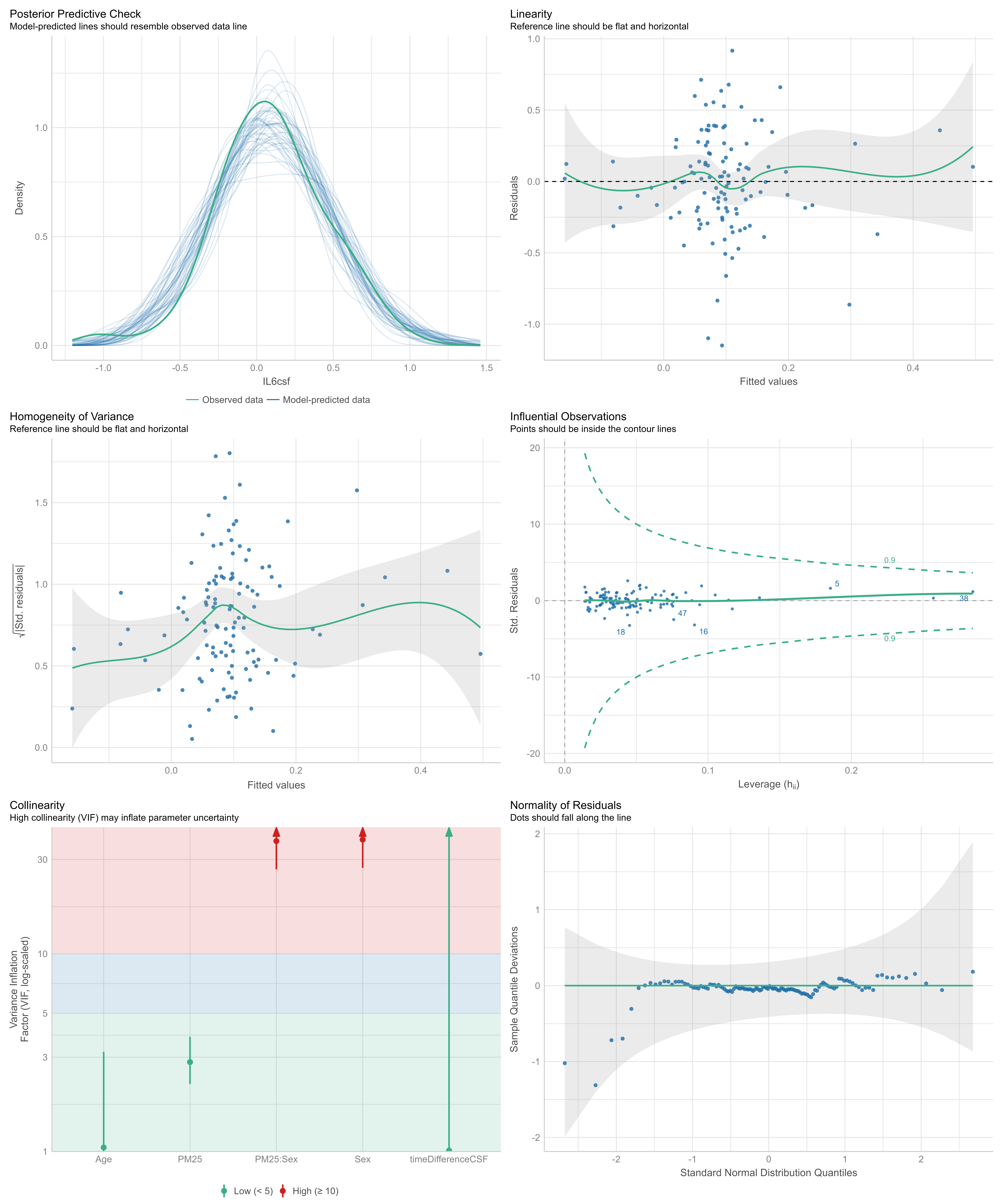

### SupplFigure10-ModelPerformance_CSFIL6_PM25abs.png

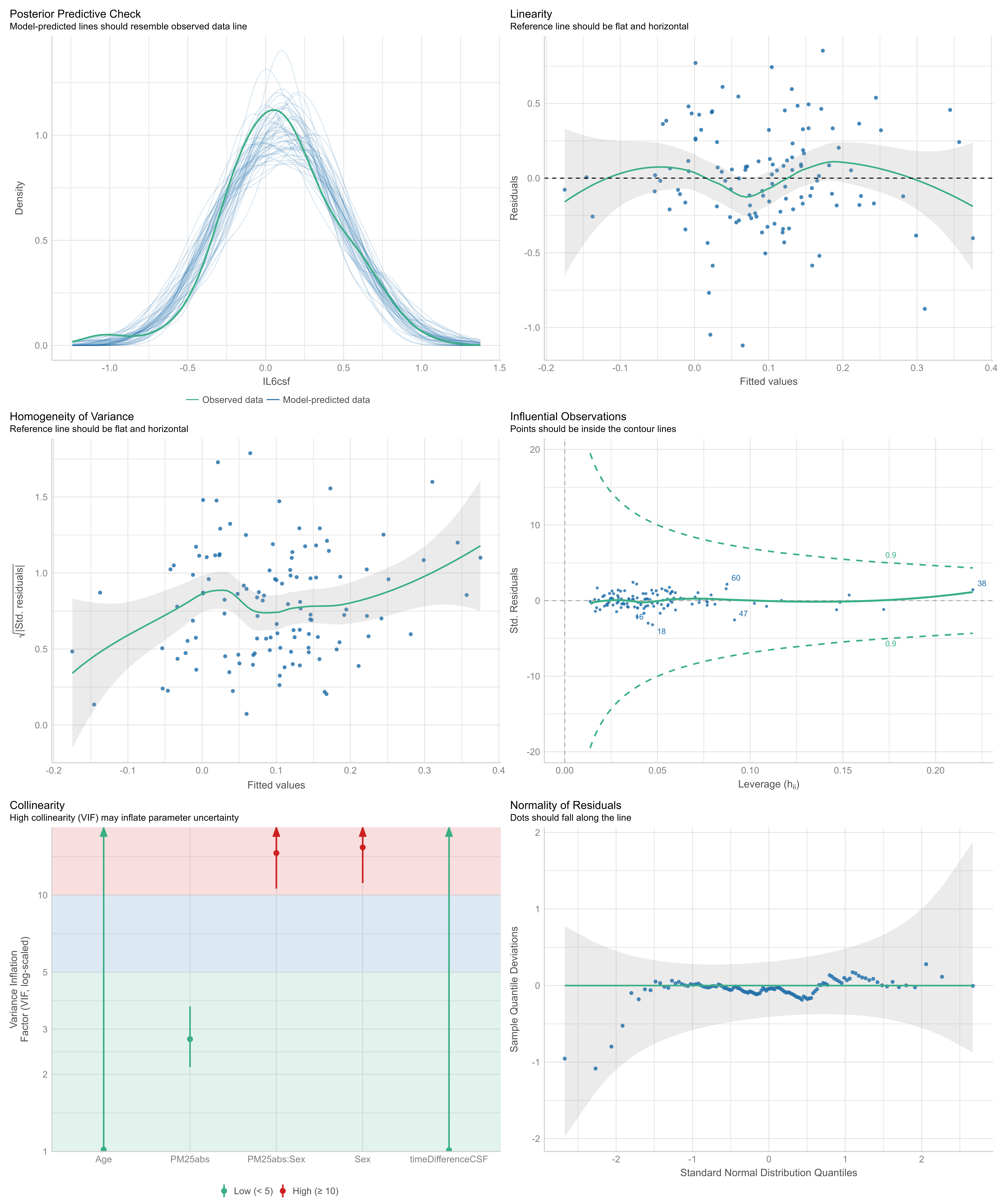
